# Implementation Science Protocol for a participatory, theory-informed implementation research programme in the context of health system strengthening in sub-Saharan Africa (ASSET-ImplementER)

**DOI:** 10.1101/2021.01.04.21249194

**Authors:** Nadine Seward, Jamie Murdoch, Charlotte Hanlon, Ricardo Araya, Wei Gao, Richard Harding, Crick Lund, Saba Hinrichs-Krapels, Rosie Mayston, Muralikrishnan R. Kartha, Martin Prince, Jane Sandall, Graham Thornicroft, Ruth Verhey, Nick Sevdalis

## Abstract

**Background:** ASSET (Health System Strengthening in Sub-Saharan Africa) is a health system strengthening (HSS) programme that aims to develop and evaluate effective and sustainable solutions that support high-quality care that involve eight work packages across four sub-Saharan African countries. Here we present the protocol for the implementation science (IS) theme within ASSET that aims to (1) understand what HSS interventions work, for whom and how; and (2) how implementation science methodologies can be adapted to improve the design and evaluation of HSS interventions within resource-poor contexts.

**Methods:** *Pre-implementation phase:* The IS theme, jointly with ASSET work-packages, applies IS determinant frameworks to identify factors that influence the effectiveness of delivering evidence-informed care. Determinants are used to select a set of HSS interventions for further evaluation, where work packages also theorise selective mechanisms to achieve the expected outcomes.

*Piloting phase and rolling implementation phase:* Work-packages pilot the HSS interventions. An iterative process then begins involving evaluation, refection and adaptation. Throughout this phase, IS determinant frameworks are applied to monitor and identify barriers and enablers to implementation in a series of workshops, surveys and interviews. Selective mechanisms of action are also investigated. In a final workshop, ASSET teams come together, to reflect and explore the utility of the selected IS methods and provide suggestions for future use. Structured templates are used to organise and analyse common and heterogeneous patterns across work-packages. Qualitative data are analysed using thematic analysis and quantitative data is analysed using means and proportions.

**Conclusions:** We use a novel combination of implementation science methods at a programmatic level to facilitate comparisons of determinants and mechanisms that influence the effectiveness of HSS interventions in achieving implementation outcomes across different contexts. The study will also contribute conceptual development and clarification at the underdeveloped interface of implementation science, HSS and global health.

**Strengths and limitations of this study:**

- The purpose of this protocol paper is to describe the methodology for the implementation science theme within ASSET (ASSET-ImplementER).
- The overall aim of the ASSET-ImplementER theme is to advance our understanding of how to design and evaluate HSS interventions using a systems level approach informed by implementation science, across different health systems and contexts.
- This is one of the first large-scale implementation research programmes for health system strengthening in Sub-Saharan Africa that spanning three care platforms (primary care for the integrated treatment of chronic conditions in adults, maternal and newborn care, surgical care), involving eight work packages in four sub-Saharan African countries that aims to apply robust, implementation science and other theory-informed approaches to understand what works for whom and how. Equally as important, we aim to improve implementation science methodologies to design and evaluate HSS interventions within LMIC settings.
- Although our programme is theory-informed through the use of ToC workshops and
- implementation science determinant frameworks, we have not used implementation science theories or other middle-range theories to guide the design and evaluate of ASSET as a programme. This is a pragmatic approach given many of the work-packages teams were unfamiliar with implementation science methods at the time of ASSET set-up.
- To mitigate the effects of this approach to our research, we work with the different work packages throughout the different phases of research, to theorise and conceptualise how the selected determinants interact with mechanisms introduced by the selected HSS on implementation outcomes.
- Although there are limitations to our approach, we expect to begin advance our understanding of what HSS work for whom, and how. It is also hoped that we will start to address the issues in understanding the complexity surrounding how to effectively strengthen health systems in resource-poor contexts within LMICs.

## BACKGROUND

Achieving Universal Health Coverage that includes the availability and delivery of high-quality evidence-informed care has been identified as an urgent priority for health system strengthening in low- and middle-income countries (LMICs).(1, 2) The scale of unmet need is reflected in estimates that that suggest 5 million people in LMICs who engaged with the health system died due to poor-quality health care in 2016, and that 3.6 million deaths occurred due to people not accessing care at all.(3) Improving population health will require not only increased access to health services, but also the provision of high-quality care.(4)

There is lack of consensus as to what constitutes health systems strengthening (HSS).(5) Initially, the term HSS was a reaction to the limitations of vertical programmes targeting specific diseases, that do not strengthen the broader health system.(5) Over time, the definition of HSS changed to one that has system-level effects and not just organisational level effects. It is now recognised that health systems are dynamic and complex socio-technical structures, composed of multiple interacting components that are constantly adapting to changes in the local context and therefore behave unpredictably.(6) HSS to improve quality of care is about “*permanently making the system function better, and not just about filling gaps or supporting the system to produce better short-term outcomes*”.(7) HSS involves comprehensive changes to policies and regulations, organisational structures, and relationships across the health system building blocks (e. g. service delivery, health workforce, health information systems, access to essential medicines, financing, and leadership/governance) that motivate changes in behaviour, and/or allow more effective use of resources across multiple care platforms.(6, 8)

Interventions to strengthen health systems, by their very nature, improve health outcomes by providing components that influence several mechanisms, both simultaneously and in isolation, at various time points and at various levels of the health system.(6) As such, HSS requires an approach to design and evaluate complex public health interventions, in real-world contexts, that accounts for the multiple interconnecting components and actors.(9) A systems-level approach to the design and evaluation of HSS interventions views a complex intervention as a system in itself, interacting with other building blocks of the underlying health system in which the intervention embeds itself, setting off reactions that may well be unexpected or unpredictable.(6) Applying this approach to the design and evaluation of HSS interventions requires an evaluation of not only their main effects, but also inputs, outputs, initial, intermediate and eventual outcomes, feedback processes and contexts within the underlying health system.(6)

### Applying implementation science concepts and methods to health system strengthening

Implementation research is a rapidly expanding discipline that seeks to understand what, why, and how interventions work in real-world settings.(10) Implementation research can consider any aspect of implementation, including understanding contextual and behavioural barriers that influence implementation efforts, the process of implementation, and the evaluation of implementation efforts for outcomes such as reach, fidelity and sustainability.(10, 11) By addressing implementation challenges through the participation of actors from multiple disciplines, implementation research can help to apply a systems-level approach and extract lessons that contribute to building stronger and more resilient and responsive health systems.(12)

Whereas implementation *research* seeks to understand what, why, and how interventions work in real-world settings,(10) implementation science is the study of methods used to carry out this research.(13) Specifically, implementation science offers theories, models, frameworks and other methodologies to optimise and evaluate the implementation of evidence-informed care.(13) The methods offered through implementation science can inform decision-makers on how best to design, implement and adapt HSS interventions in the dynamic environments in which they work, to produce new knowledge for more effective strategies and implementation.(14) For example, implementation science determinant frameworks describe contextual and behavioural barriers that are known to influence implementation outcomes, and include key factors to consider in the evaluation of the process of implementation.(15) Box 1 describes key implementation science definitions as used within the ASSET-ImplementER theme.

#### Box 1

**Implementation science terminology used within the ASSET-ImplemenER programme**

##### Implementation strategies

methods or techniques used to enhance the adoption, implementation and sustainability of a clinical program or practice. Other terminology includes HSS interventions, quality improvement strategies.

##### Implementation outcomes

Implementation outcomes assess how well implementation has occurred or provide insights about how this contributes to an individual’s health status or other important health outcomes. Proctor et al articulate the following core set of seven implementation outcomes: acceptability, feasibility, uptake, penetration, cost, fidelity, and sustainability.(16)

##### Implementation effectiveness

the impact of implementation efforts on implementation outcomes.

##### Evaluation framework

Specifies implementation outcomes that can be evaluated to determine implementation effectiveness.(17)

##### Context

Any feature of the circumstances in which an intervention is conceived, developed, implemented and evaluated.(18) Other commonly termed setting or environment.

##### Determinant frameworks

identify contextual barriers and/or enablers that are known to impact on the effectiveness of implementation efforts.(15)

##### Theoretical frameworks

identify determinants of behaviours that are known to influence implementation outcomes.(19)

##### Implementation theories/middle range theories

describe the mechanisms behind how a proposed intervention works. These theories can also be used to identify barriers and/or enablers to change and what needs to change.(15)

##### Programme theory

describes how a specific intervention is expected to lead to its effects and under what conditions.(20)

### Need for improved methods to design and evaluate health system strengthening interventions in LMICs

Despite the need to account for the complexity of HSS interventions, current methods used to design and evaluate such interventions in LMICs typically lack robust methodologies that enable an understanding of what interventions worked, for whom, and how.(21) As an example, few studies to-date have been supported by a conceptual framework (including a programme theory) explaining how an intervention is theorised to work, or a detailed process evaluation that helps to explain whether the intervention worked as intended, and if so, for whom, and under what circumstances.(21-24) There are also limitations with how studies report how the local context influences the effectiveness of HSS interventions on important implementation outcomes such as coverage, acceptability and fidelity. Importantly, much of the evidence base for HSS interventions still comes from high-income countries, with uncertain generalisability of evidence to low-resourced settings.(25)

Compounding the methodological shortcoming with HSS in LMICs, are the inconsistences in the terminology used to describe the science of promoting and supporting the use of evidence in health and healthcare policy(26, 27) and also its reporting. As an example, implementation science has been described variously as quality improvement, knowledge translation, knowledge transfer, and even complex intervention evaluation.(27) Although there are differences between fields such implementation science and improvement science, they share the objective of improving the quality of healthcare delivery and therefore clinical outcomes.(28, 29)

Perhaps even more important is the lack of consistency in reporting and describing the implementation strategies to improve the quality of service delivery and therefore clinical outcomes.(30, 31) This results in the inability to replicate or generalise research studies to different contexts, or to allow for research synthesis such as meta-analysis or systematic reviews.(32, 33) To help address this issue, the Expert Recommendations for Implementing Change (ERIC) study created a taxonomy of implementation strategies(32) – to allow researchers to apply a common language when describing how evidenced interventions are being implemented. Taxonomies for quality improvement strategies have also been created that are conceptually similar (in some cases identical) to the ERIC taxonomy.(32, 34) As an example, both taxonomies include audit and feedback strategies, provider education, patient education, patient reminders and organisational change.(32, 34) There is also a taxonomy for HSS interventions, the Effective Practice and Organisation of Care (EPOC) taxonomy, that is also similar to the taxonomies for implementation and quality improvement strategies.(32, 35)

There is, therefore, an urgent need to improve not only the quality of implementation research for HSS in LMICs, but also consistency in reporting the implementation strategies used to deliver evidence-informed care and improve service delivery and patient outcomes. For purposes of the ASSET-ImplementER theme, we will refer to these interventions as HSS interventions. Embedding high-quality implementation science methodology in HSS research that includes consistent reporting of the HSS interventions used to deliver evidence-informed care, will help unpack the ‘black box’ of how such interventions work (or fail to reach their expected potential) for certain populations in a given setting.(22) The ASSET-ImplementER study aims to start addressing some of these issues.

### The National Health Institute of Research (NIHR) Global Research Unit on Health System Strengthening in Sub-Saharan Africa (ASSET) and the ASSET-Implementation Research theme (ASSET-ImplementER)

ASSET is a four-year HSS research programme (2017-21) that addresses the imperative of using implementation research with robust implementation science methods that use consistent reporting of the HSS interventions to deliver high-quality care. ASSET spans three healthcare platforms (primary health care for the integrated treatment of chronic conditions in adults, maternal and newborn, surgical care) involving eight work packages within four different countries within sub-Saharan Africa. The overall aim of ASSET is to develop, implement and evaluate high quality HSS interventions that are effective and sustainable. ASSET also aims to ensure care is equitable, people-centred and respectful.(36) ASSET is one of the first implementation research programmes for HSS that applies an ‘effectiveness-implementation hybrid’ approach, blending components of study designs that evaluate the effectiveness of interventions on patient outcomes, with study designs that focus on evaluating the effectiveness of HSS interventions on implementation outcomes.(37) This methodology allows ASSET to understand both clinical and implementation effectiveness as well as contextual factors influencing implementation outcomes, including the potential sustainability of the different HSS interventions.

The ASSET programme includes two phases of implementation research including the pre-implementation phase, and piloting and rolling implementation phase. The *pre-implementation phase* aims to understand requirements for HSS through the evaluation of the structure and function of the health system (such as arrangements for financing, governing, and delivering care, and implementation strategies) (35) that may limit the potential to deliver evidence-informed and person-centred care. At the end of this phase, each work package use research findings to select a set of HSS interventions. The *piloting and rolling implementation phase initially* pilot the interventions to identify factors that influence the implementation of the proposed interventions. After adjusting the initial programme theory and set of HSS interventions, an adaptive and iterative process is used to test the effectiveness of the set of HSS interventions on both clinical and implementation outcomes. The influence of context on the effectiveness of the HSS interventions delivering evidence-informed care is also assessed.(36)

### Aim and objectives

The overall aim of the ASSET-ImplementER theme is to advance our understanding of how to design and evaluate HSS interventions using a systems level approach informed by implementation science, across different health systems and contexts. In doing so, we expect to achieve two equally important objectives for HSS in resource-poor contexts in LMICs: (1) advance our understanding of what HSS interventions work, for whom and how; and (2) improve implementation science methodologies to design and evaluate HSS interventions within LMIC settings.

The following specific objectives will be addressed within the ASSET-ImplementER theme

### Pre-implementation and piloting phases

1. Contrast and compare the contextual determinants identified by each work package that influence the effectiveness of evidence-informed care;
2. Contrast and compare HSS interventions selected by each work package that have the potential to improve the effectiveness of evidence-informed care;
3. Determine how the identified contextual determinants influence ASSET investigators in selecting specific HSS interventions and associated implementation outcomes and compare findings between ASSET work-packages.

### Implementation and evaluation phase of ASSET HSS interventions

4. Contrast and compare findings between the different work-packages on how context influences the delivery of the selected HSS interventions on standardised implementation outcomes measures for acceptability, appropriateness and feasibility;
5. Evaluate the usefulness and utility of the selected implementation science methods/frameworks in achieving a systems level approach in the evaluation of HSS interventions in resource poor contexts and provide suggestions for future applications of implementation science to HSS.

## METHODS

### Setting

ASSET is working on three care platforms: (1) surgical care; (2) maternal and newborn care, and; (3) integrated primary care, across four sub-Saharan African countries: Ethiopia, Sierra Leone, South Africa, and Zimbabwe. Table 1 describes the ASSET eight work-packages.

**Table 1:**
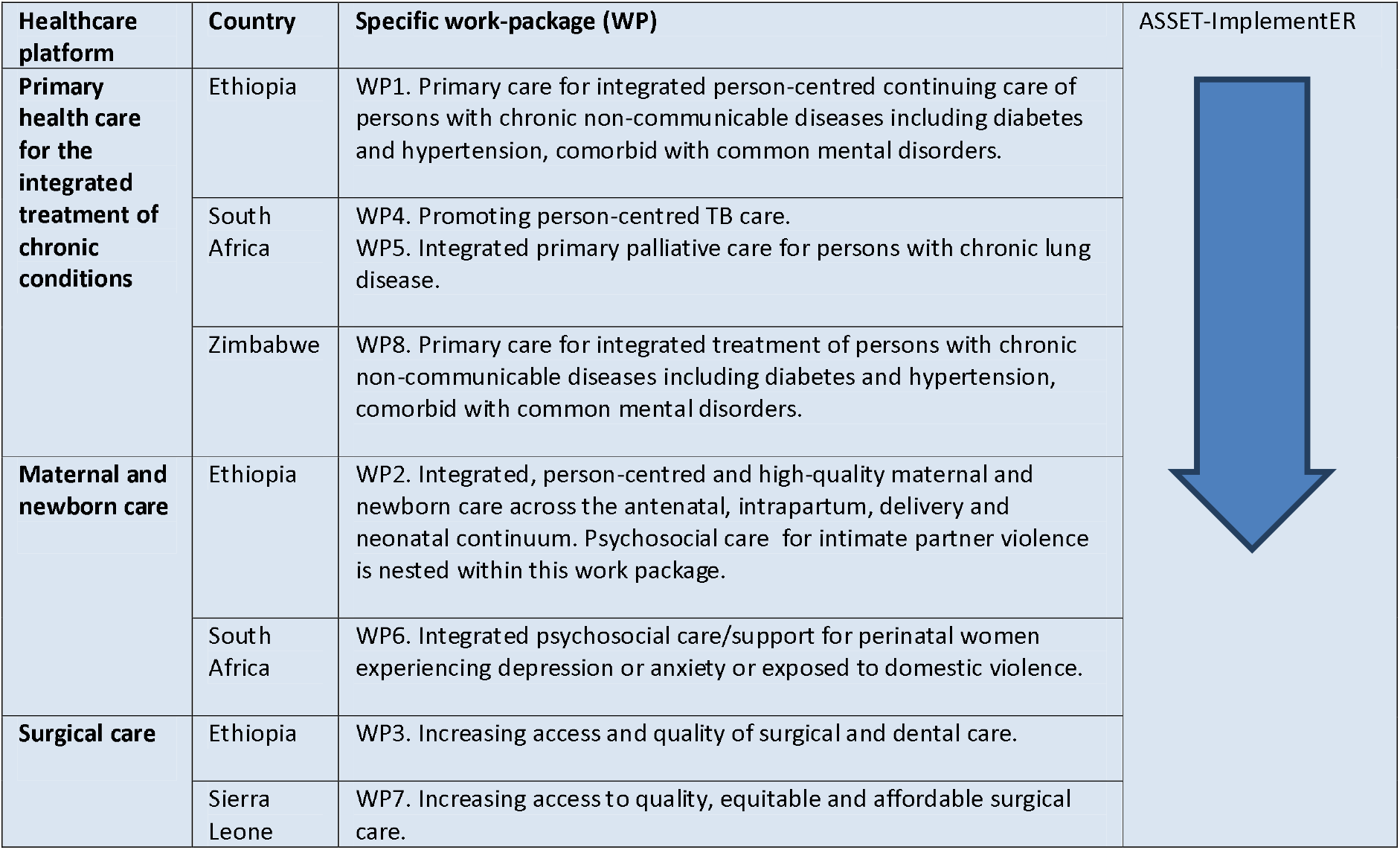
Description of the ASSET work-packages for the different healthcare platforms; implementation research cuts across all of them (ASSET-ImplementER theme)

| Healthcare platform | Country | Specific work-package (WP) | ASSET-ImplementER |
| --- | --- | --- | --- |
| **Primary health care for the integrated treatment of chronic conditions** | Ethiopia | WP1. Primary care for integrated person-centred continuing care of persons with chronic non-communicable diseases including diabetes and hypertension, comorbid with common mental disorders. |  |
|  | South Africa | WP4. Promoting person-centred TB care.<br>WP5. Integrated primary palliative care for persons with chronic lung disease. |  |
|  | Zimbabwe | WP8. Primary care for integrated treatment of persons with chronic non-communicable diseases including diabetes and hypertension, comorbid with common mental disorders. |  |
| **Maternal and newborn care** | Ethiopia | WP2. Integrated, person-centred and high-quality maternal and newborn care across the antenatal, intrapartum, delivery and neonatal continuum. Psychosocial care for intimate partner violence is nested within this work package. |  |
|  | South Africa | WP6. Integrated psychosocial care/support for perinatal women experiencing depression or anxiety or exposed to domestic violence. |  |
| **Surgical care** | Ethiopia | WP3. Increasing access and quality of surgical and dental care. |  |
|  | Sierra Leone | WP7. Increasing access to quality, equitable and affordable surgical care. |  |

### Design

The work-packages use mixed-methods throughout ASSET to select HSS interventions in the pre-implementation phase and evaluate the intervention through ‘effectiveness-implementation hybrid’ designs in the piloting and rolling implementation phase. Hybrid designs are essential with implementation research as they blend the components of study designs used to evaluate clinical effectiveness, with those of implementation study designs that focus on the evaluation of the influence of context on the effectiveness of HSS interventions.(37)

Throughout ASSET, the ASSET-ImplementER stream uses mixed methods including workshops, semi-structured interviews and documentary analyses, to standardise, record and synthesise findings from the implementation component of the different work-packages. Findings include information from the different frameworks such as context, intervention, selected HSS interventions, and implementation outcomes. Figure 1 describes the flow of methods for the ASSET-ImplementER theme.

**1.**
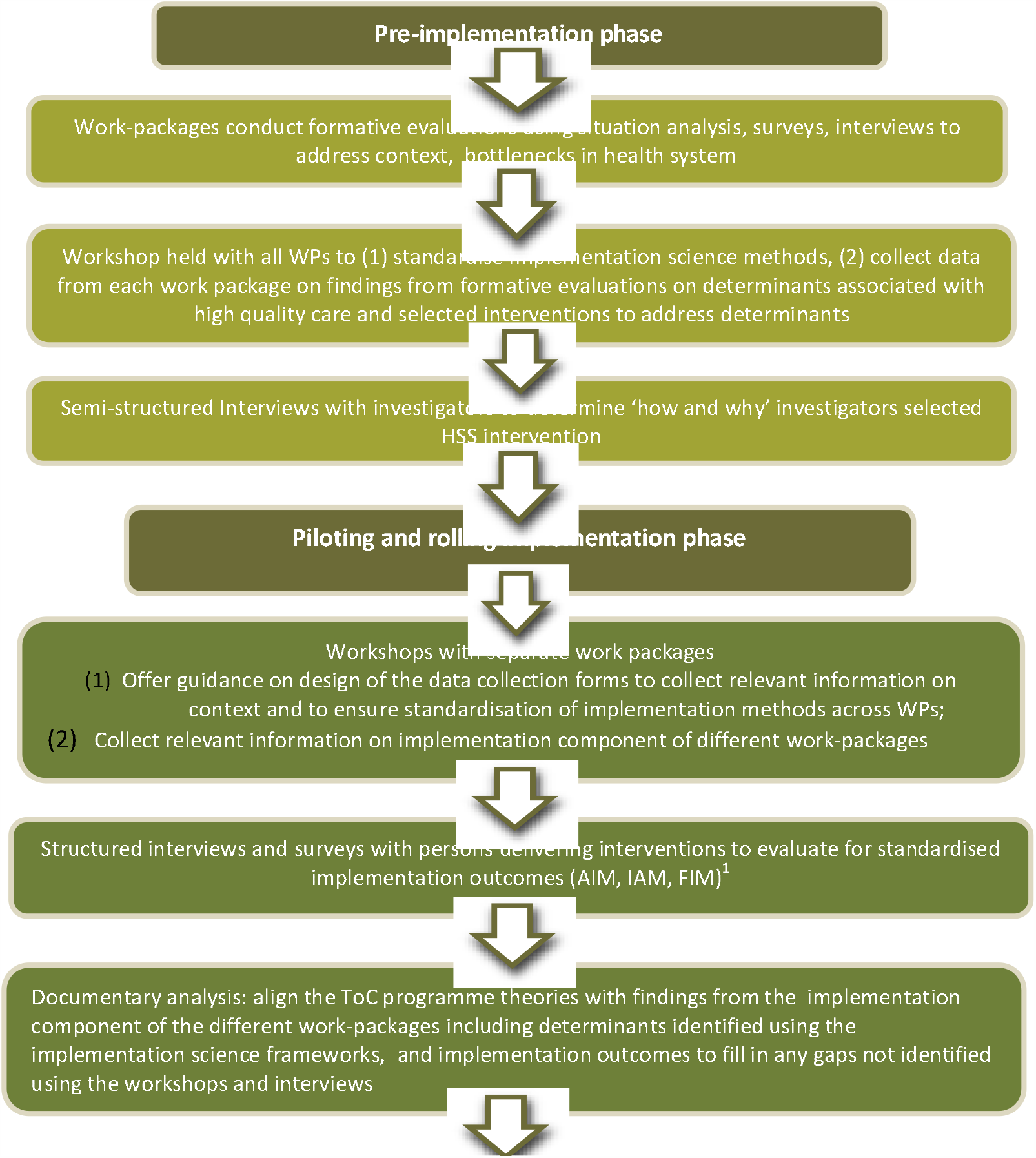
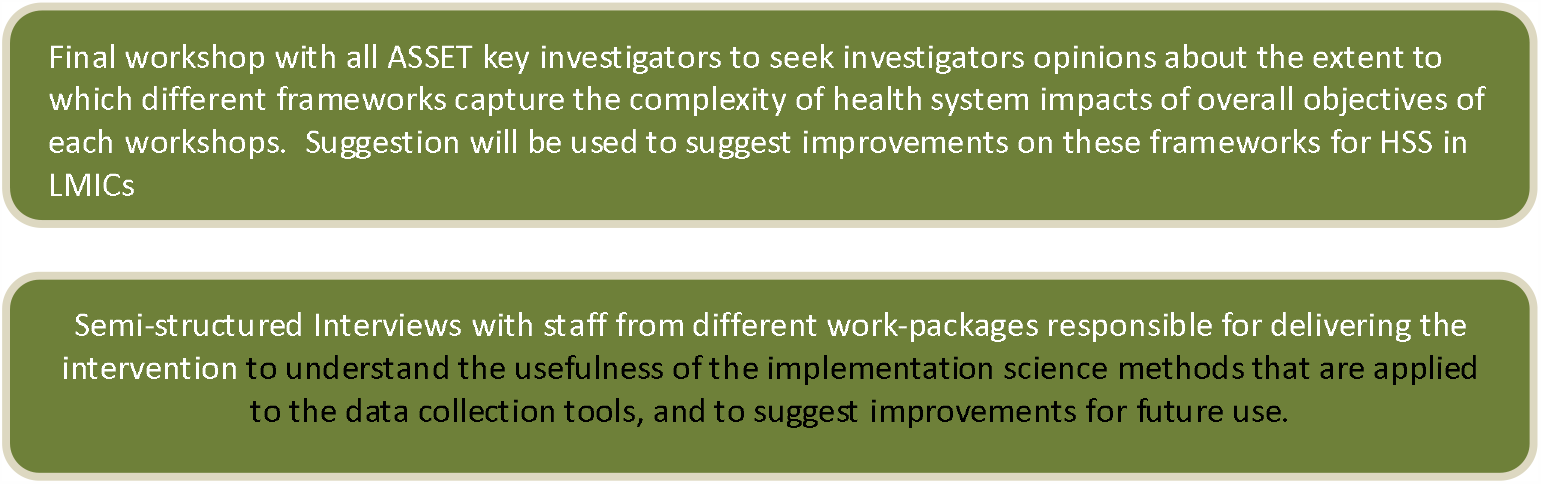
Acceptability of Intervention Measure (AIM), Intervention Appropriateness Measure (IAM), and Feasibility of Intervention Measure (FIM), have demonstrated promising psychometric properties in high-income settings.(38)

**Figure 2: Flow of methods for the ASSET-ImplementER theme**

To standardise methods and facilitate cross-site comparisons across ASSET, work-packages select contextual and behavioural determinants and implementation outcomes from a defined set implementation science frameworks. These frameworks are relevant to the programme as a whole, yet at the same time account for the specific characteristics of the different work-packages. Table 2 describes the frameworks that the various work-packages use and how this is relevant to the overall ASSET programme.

**Table 2:** Selected implementation science frameworks and theories used within ASSET-ImplementER.

| Framework | Framework type and description | Relevance to ASSET |
| --- | --- | --- |
| **Consolidated framework for Implementation Research (CFIR)** | The CFIR is a determinant framework that Includes five domains (inner setting, outer setting, intervention characteristics, characteristics of individuals involved, and the processes of implementation).(39) Within the five domains, are 37 constructs that can behave as a barrier and/or enabler to the implementation of an intervention | The appeal of the CFIR framework is the broad range of constructs that apply not only to individual characteristics, but also structural characteristics of the health care system as well as characteristics of the intervention itself. (39) |
| **Context and Implementation of Complex Interventions (CICI)** | The CICI is both a determinant and evaluation framework that contains three dimensions (context, implementation and setting) that interact with the intervention. The contextual dimension consists of seven contextual domains (i.e. geographical, epidemiological, socio-cultural, socio-economic, ethical, legal, political) with interact with one another at the micro, meso and macro levels.(40) | The appeal of this framework is the emphasis on the external contextual elements such as sociocultural, socioeconomic and political determinants. These determinants are particularly relevant in low-resource contexts.<br>This framework also explicitly addresses complexity required with the design and evaluation of HSS interventions, |
|  |  | through the evaluation of the interaction of context (including leadership elements), implementation and setting dimensions with the intervention dimension, at the micro, meso, and macro levels. |
| **Theoretical Domains Framework (TDF)** | The implementation of evidence-informed interventions is dependent on changing multiple behaviours of different people.(41) where the use of theory to inform behaviour change interventions has been shown to improve the implementation effectiveness.(42) We will adopt the Theoretical Domains Framework (TDF) that represents a synthesis of 128 determinants of behavioural change.(19) | Many of the barriers to HSS interventions for ASSET are associated with determinants of current and desired behaviours e.g. in health worker or health service users. As an example, social norms that stigmatise common mental health conditions can act as a barrier to respectful, person-centred care. These determinants are particularly relevant to providing person-centred respectful care as well as high quality care. |
| **Behaviour Change Wheel** | The implementation of evidence-informed practice is dependent on effective behaviour change interventions. The Behavioural Change Wheel is framework that includes nine intervention functions that address determinants of problematic behaviours that require changing. | To ensure longer term sustainability, implementing evidence-informed practice for HSS will involve a degree of behavioural change. Once determinants of the problematic or desirable behaviours have been identified using the TDF, the BCW can be applied to help select appropriate interventions that have demonstrated effectiveness. |
| **Proctor et al Implementation outcome framework** | An evaluation framework that includes the following eight implementation outcomes: acceptability, adaptability, appropriateness, feasibility, fidelity, cost, penetration, and sustainability.(43) | All of these outcomes are relevant to ASSET at different time points in the implementation of the HSS interventions. As an example, the acceptability, appropriateness and feasibility of the HSS interventions are important to measure in the pre-implementation/piloting phases as they can predict the longer-term sustainability of the intervention. (ref) |

The Consolidated Framework of Implementation Research (CFIR) is a determinant framework that is used as it provides an overview of a broad range of determinants that influence implementation effectiveness such as the inner setting (i.e. characteristics of the health facility), characteristics of the intervention (e.g. acceptability, complexity and adaptability) and implementation processes (e.g. regular feedback about progress and quality of implementation).(39) The Context and Implementation of Complex Intervention (CICI) framework is another framework that we use as it offers a detailed approach to identifying determinants from the external context (e.g. sociocultural, socioeconomic, political, epidemiological, ethical, and legal) that are known to influence implementation effectiveness that are particularly relevant to LMICs.(40) It is expected that both of these frameworks will provide a detailed spectrum of determinants that are relevant to the ASSET programme. However, the frameworks are not exhaustive: any determinant identified that is not a part of either frameworks will be documented as such.

Further to contextual determinants, it is also important to understand in some detail characteristics of the users who deliver the healthcare. Changing behaviour ingrained in individuals working within health systems will help to ensure the adoption longer term sustainability of the HSS interventions. To address this, we will use the Theoretical Domains Framework (TDF), a determinant framework that brings together evidence based determinants of behaviour.(19) We also use the Behaviour Change Wheel (BCW), that explicitly maps behavioural change interventions onto determinants of behaviour identified with the TDF.(44) Each work-package selects the contextual and behavioural determinants, implementation outcomes and HSS interventions from the different frameworks that are relevant to their aims and objectives. Table 3 describes the objectives of the different implementation science frameworks across the different phases of ASSET. However, identifying determinants that influence the effectiveness of HSS interventions in delivering evidence-informed practices is not enough to address complexity associated with HSS. We will therefore also explore how context influences the mechanisms introduced by the HSS interventions on implementation outcomes.

**Table 3:**
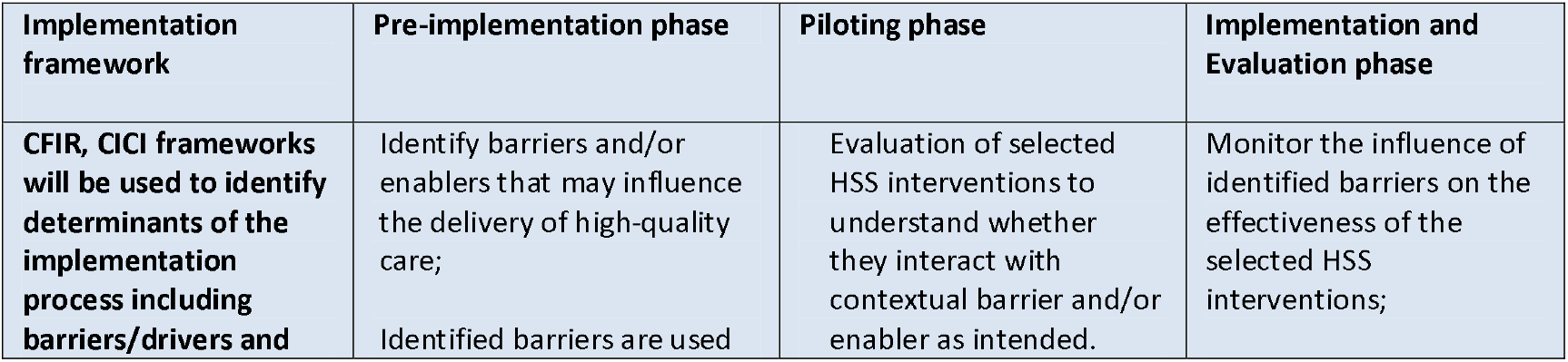
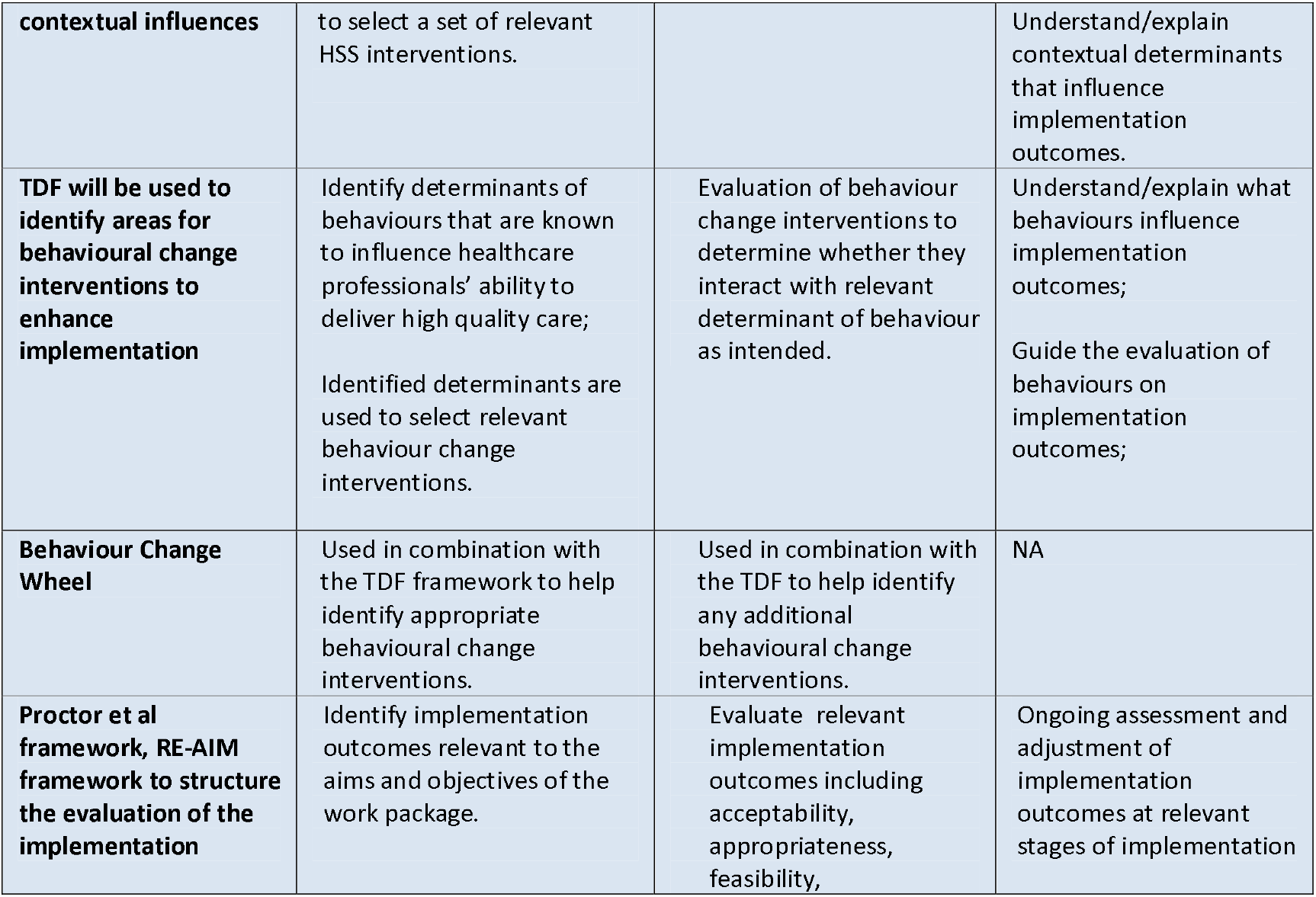
Application of implementation science frameworks across ASSET health system strengthening interventions and research phases.

### Measures and taxonomies

#### Implementation outcomes

To evaluate the effectiveness of the implementation efforts, ASSET uses a combination of implementation outcomes as defined by different evaluation frameworks.(16, 17) The selected implementation outcomes are aligned with the aims and objectives for the different work-packages.

Many measures used to evaluate implementation outcomes have not been validated, making it difficult to compare the effectiveness of alternative interventions both within and between studies.(38) However, three implementation outcomes measures including Acceptability of Intervention Measure (AIM), Intervention Appropriateness Measure (IAM), and Feasibility of Intervention Measure (FIM), have demonstrated promising psychometric properties in high-income settings.(38) Work-packages test the applicability of these measures in low-resourced settings by adapting them to their specific context. Each measure includes four questions measured on a Likert scale (completely disagree, disagree, neither agree nor disagree, agree, completely agree). Questions for each measure will be translated and back translated to ensure linguistic and cross-cultural equivalence. The questions will then be reviewed and cognitive testing with local participants performed prior to the tools being used.

#### Taxonomies

Initially we use the EPOC taxonomy for HSS interventions to label the different interventions selected by work-packages.(35) If an appropriate intervention is not included in this taxonomy, work-packages use the ERIC taxonomy for implementation strategies to label the intervention.(32) To help alleviate the ambiguity in reporting implementation strategies and HSS interventions, we also report any important differences and similarities between the two taxonomies.

To provide consistency in labelling of the behavioural change interventions, we use the taxonomy of behavioural change interventions that is also aligned with the TDF and the BCW.(45) However, there are important limitations to applying the TDF and BCW to help select interventions that improve the quality of people-centred care that also address issues such as illness-related stigma. As an example, these behavioural change methods were not specifically designed to address the issues surrounding implementation research in low-resource setting such as stigma, that ASSET is focusing on. As such the behavioural change interventions in this taxonomy may not be appropriate. To address this issue and similar issues with the other taxonomies, where the HSS interventions, quality improvement strategy or behavioural change intervention is not described in any taxonomy, we clearly describe the intervention and highlight the issue with the selected taxonomy not describing our intervention.

#### Reporting frameworks

To ensure consistent and accurate reporting of implementation studies, work-packages will apply the Standards for Reporting Implementation Studies (StaRI).(46)

## Data collection procedures

### Data collection in the pre-implementation phase

#### Workshops

At the end of the pre-implementation phase of ASSET, a two-day workshop is held involving relevant participants from all work-packages (i.e. principal investigators, co-investigators, field staff, and research assistants). The objectives of the workshop are to: (1) standardise the implementation methods being applied across the different work-packages, and; (2) collect data from each work-package on contextual and behavioural determinants associated with the delivery of high-quality evidence-informed care and the associated HSS interventions and behavioural change interventions that were selected to help deliver this care.

Using data collection tools that identify and describe determinants of the implementation process, participants within the different work-packages apply findings from the formative research to identify and record barriers and/or enablers to the delivery of high-quality care. Participants then use the selected determinants to theorise how potential HSS interventions described in the EPOC taxonomy, can assist in delivering high-quality evidence-informed care relevant to their work package. Similar data collection methods and forms are applied to label and record determinants of behaviours that influence the delivery of evidence-informed care. The identified determinants are then be used to select and record behavioural change interventions.

#### Interviews & focus groups

The workshop is followed-up with semi-structured virtual interviews involving investigators from the different work-packages, to help understand the ‘how and why’, which led them to select the HSS interventions and associated strategies to facilitate their implementation (or virtual focus groups if more feasible for country teams). The same interviews/focus groups are also used to review and finalise the findings from the workshop and to understand if there are any gaps in evidence that will require the review any additional documentation. We anticipate including a minimum of two investigators from each ASSET work-package (16 investigators minimum sample) to reach saturation of the thematic areas that emerge from their responses.

### Data collection in the piloting and rolling implementation phase

#### Workshops

Throughout the piloting and rolling implementation phase, workshops are held with each work package team, involving relevant participants (principal and co-investigators, field staff, research assistants, and PhD students). The purpose of the workshops is to guide investigators on how to design the data collection tools including interview guides, using the implementation science determinant frameworks, HSS taxonomies and implementation outcomes. In doing so, we hope to ensure findings are standardised and therefore comparable across work-packages.

At the end of the piloting and rolling implementation phase, a separate workshops will be held with each of different work-package teams to feedback the following information collected using the data collection tools: (1) the effectiveness of HSS interventions on standardised implementation outcomes, (2) the influence of context on the effectiveness of HSS intervention in delivering evidence-informed care, and (3) the direct influence of context on the mechanisms introduced by the intervention to produce change.

At the end of the piloting and rolling implementation phase, another workshop lasting one day and involving investigators from all work-packages is used to reflect on the different implementation science methodologies and suggest improvements for further use. Specifically, we seek investigators’ opinions about the extent to which different frameworks capture the complexity of how the health system impacts on the overarching problem each work-package is trying to address. Again, we anticipate including a minimum of two investigators from each ASSET work-package (16 investigators minimum sample) to reach saturation of the thematic areas that emerge from their responses. All meetings will be audio-recorded with descriptive notes of discussion. Findings from these workshops are used to suggest how frameworks can be adapted to capture this complexity.

#### Surveys

In select work packages (WP 1, 2, 5, 6, 8) staff delivering the interventions (i.e. nurses, community health workers) are asked to complete the linguistically adapted tools for the three outcome measures, including acceptability (AIM), appropriateness (IAM) and feasibility (FIM).(38) Approximately 60 staff members from each of the participating work packages contribute to these surveys. A subset of these participants are then interviewed to discuss the usefulness of the data collection tools. In particular, we review methods to capture the influence of context on both the effectiveness of the HSS interventions in influencing implementation outcomes as well as the influence of context on influencing the mechanisms introduced by the HSS interventions on implementation outcomes. Participants are also questioned on recommendations to improve our ability to capture the influence of context on implementation outcomes.

#### Documentary analysis

Theory-of-Change (ToC) methodology is a participatory approach involving key stakeholders that allows the articulation of the ‘theory’ of how a complex interventional programme will work in reality, describing the necessary interventions to bring about the change, as well as the assumptions inherent to the programme and importantly the context of implementation.(47) ASSET work-packages are developing ToCs to support their implementation and evaluation planning. ToCs are effectively programme theories, contextualised within each one of the ASSET work-packages, offering an overview of how the selected HSS interventions are theorised to achieve specific implementation outcomes. ToCs also include information on key assumptions and work-package context. Each work package including all relevant stakeholders, develops and initial ToC in the pre-implementation phase. This programme theory is then adapted throughout the piloting and pre-implementation phase. We examine whether and how the ToCs align with findings from both the pre-implementation and piloting and rolling implementation phases of the different work-packages including determinants identified using the implementation science frameworks outlined in Table 3. We will also use ToC to better articulate how context influences mechanisms introduced by the HSS interventions described by each work package.

### Data Analysis

#### Psychometric assessment of outcome measures AIM, IAM and FIM

The outcome measures AIM, IAM and FIM that have demonstrated promising psychometric properties in high-income countries, are tested for similar properties including substantive and discriminant content validity (the extent to which a measure is judged to be reflective of a construct of interest) and inter-item consistency (extent to which scale items are scored in a similar manner) by the different work packages (45) to determine their relevance in low-resource settings. Measures are adapted to the local context, translated and back translated.

To test for substantive validity (extent to which a measure is reflective of the construct of interest) and discriminant content validity (extent to which a measure that are not supposed to be related are actually unrelated,) different cadres of workers who are responsible for delivering the intervention in the different work packages assign 31 items reflecting the three constructs, to each of the three constructs and rate their confidence in the assignments in order test. The Wilcoxon one-sample signed rank test or t-test (as appropriate) is used to determine whether items measured the intended construct, or whether items measured a combination of constructs. Hochberg’s correction is used to correct for multiple tests.(48) Intraclass correlation coefficients (ICCs) using a two-way mixed effects model to assess the level of agreement in item assignments among all participants, and also within key stakeholder groups, across 31 items and for each construct.(49) The same data are also used to assess the factorial validity of the three measures, initially through exploratory factor analyses.(38) We assess inter-item consistency by computing Cronbach alpha for each of the four-item scales. For each measure, we also calculate means and standard deviations. Higher scores represent more favourable responses. If the measures demonstrate adequate psychometric properties, they are used to facilitate cross-site comparisons across the different work-packages.

#### Data collected by ASSET-ImplementER

For each work package, a thematic analysis is used to analyse the qualitative data collected in semi-structured interviews and workshops that identify key issues pertinent to using implementation science frameworks and implementation outcome measures for HSS in low-resourced contexts.

#### Analysis and synthesis of data collected by different work-packages

At the end of each phase of research, data is collected from each of the work packages (i.e. contextual and behavioural determinants, mechanisms, selected HSS interventions and implementation outcomes) and entered into a template created in Excel software. As an example, each work-package records information on identified barriers/enablers, associated data source, relevant implementation science framework, EPOC HSS interventions, and implementation outcomes. Table 4 demonstrates an example of the Excel template.

**Table 4:** Sample template used to synthesise findings from the workshops and interviews.

| **Determinant** | **Data source for determinants** | **Implementation science framework** | **EPOC health system strengthening intervention** | **ERIC implementation strategy** | **Behavioural change intervention** |
| --- | --- | --- | --- | --- | --- |
| Illness-related stigma | Focus-group discussion with key stakeholders | CICI/TDF | Educational meetings with health care providers, educational materials distributed to patients and clinicians | Education and training | Motivate health workers |

We adapt the matrixed multiple case study approach to analyse and synthesise the data we record in the templates. (20) This method facilitates comparisons between relevant work-packages, organising, analysing and presenting common and heterogeneous findings across implementation sites. Such an approach also aims to create generalisable knowledge regarding what and how local factors influence implementation.(20) The matrixed multiple case study approach uses a combination of quantitative and qualitative methods that will allow us to identify associations between specific implementation processes and contextual factors on the one hand, and implementation outcomes on the other.

Initially, data are analysed separately for each work package. Quantitative data are analysed using means and proportions. Given the heterogeneous nature of the care platforms and associated work-packages, context, and selected HSS interventions, it will not be useful to compare quantitatively the influence of context on the effectiveness of HSS interventions on implementation outcomes between the work-packages. Instead, we use a qualitative approach that aims to understand why implementation outcomes were similar or different by describing the associated HSS interventions and contextual and behavioural determinants.

Results from the matrixed multiple case study approach as well as findings from the interviews and the final workshop will be triangulated to identify points of convergence and divergence between different work-packages. Using these methods, we analyse how each work-package defined and operationalised different implementation science constructs (e.g. context, intervention mechanisms, HSS interventions, behaviour change techniques), and how implementation theories and frameworks were used to support specific HSS interventions throughout implementation. The final part of our analysis integrates these elements to offer an overarching understanding of how implementation science frameworks have been operationalised across all ASSET work-packages and whether the use of these frameworks offered added utility (from a design, implementation or evaluation perspectives).

### Patient public involvement

Patients and the public were not involved in the designing/writing protocol for this protocol. However, extensive participatory methods that involve both the patients and public will be used by the individual work packages to design, select and evaluate the HSSIs for ASSET.

### Ethics

The ASSET-ImplementER theme is considered minimal risk as we only interview researchers involved in the different work packages. To this effect we have received approval from King’s College London Ethics Committee for research that is considered minimal risk (Reference number: MRA-20/21-21772).

## Discussion

To our knowledge, ASSET-ImplementER is one of the first global health implementation research programmes that attempts to standardise methodologies to design and evaluate HSS interventions across different healthcare platforms and settings. In doing so, we expect to improve our understanding of what HSS interventions work for whom, by creating a compilation of HSS interventions used by the different work-packages, the associated barriers, and the effectiveness on implementation outcomes.

ASSET-ImplementER will also attempt to develop novel insights into how we can improve implementation science methodologies for designing implementation research on HSS in LMICs. Importantly, there is a lack of standardisation in not only the terminology used to describe implementation research and HSS, but also the methods applied to this research. By labelling contextual and behavioural determinants using established frameworks and testing psychometric properties of implementation outcome measures that have demonstrated similar properties in high-income countries, we hope to improve our ability to compare the effectiveness of different HSS interventions. Importantly, many researchers find the plethora of implementation science frameworks and theories overwhelming and difficult to apply in practice. Through interviews with different stakeholders, we hope to improve methods for HSS in LMICs including our ability to learn how to best apply and simplify frameworks across different health systems.

There are limitations to ASSET-ImplementER. Although our programme is theory-informed through the use of ToC workshops and implementation science determinant frameworks, we have not used implementation science theories or other middle-range theories to guide the design of ASSET as a programme. Indeed, many people criticise determinant frameworks (which we apply heavily within ASSET) as being a theoretical. Nevertheless, given many of the work-packages teams were unfamiliar with implementation science methods at the time of ASSET set-up, we feel this is a pragmatic approach to applying a theory-informed approach to HSS. To mitigate the effects of this static approach to our research, we work with the different work packages throughout the different phases of research, to theorise and conceptualise how the selected determinants interact with mechanisms introduced by the selected HSS on implementation outcomes.

ASSET-ImplementER applies a system-level approach to both the design and evaluation of HSS interventions for the ASSET programme. Although there are limitations to our approach, we expect to begin advance our understanding of what HSS work for whom, and how. It is also hoped that we will start to address the issues in understanding the complexity surrounding how to effectively strengthen health systems in resource-poor contexts within LMICs.

## Data Availability

This is a protocol paper and therefore no data is available

## Abbreviations

ASSET: Global Research Unit on Health System Strengthening in sub-Saharan Africa
AIM: Acceptability of Intervention Measure
ASSET-ImplementER: Implementation science theme within ASSET
BCW: Behavioural Change Wheel
CFIR framework: Consolidated Framework for Implementation Research
CICI framework: Context and Implementation of Complex Interventions
EPOC taxonomy: Effective Practice and Organisation of Care taxonomy for health system strengthening interventions
ERIC taxonomy: Expert Recommendations for Implementing Change taxonomy of implementation strategies
FIM: Feasibility of Intervention Measure
HSS: Health systems strengthening
IAM: Intervention appropriateness Measure
LMICs: Low- and Middle-Income Countries
NIHR: National Health Institute of Research
STaRI: Standards for Reporting Implementation Studies
TDF: Theoretical Domains Framework
ToC: Theory of Change
WP: Work package

## Declarations

### Patient consent

Protocol paper so consent not applicable

### Data sharing statement

No additional data available as a protocol paper

### Ethics

Approval from King’s College London Ethics Committee for research that is considered minimal risk (Reference number: MRA-20/21-21772).

## Authors contributions

NSeward wrote the first and subsequent drafts of the paper

NSeward and NSevdalis conceptualised the idea for the paper

NSeward, NSevdalis, Jamie Murdoch and Charlotte Hanlon offered insight and edited all drafts of the paper

All other authors (RA, WG, RH, CL, SHK, RM, MK, MP, JS, AS, GT, RV) edited and offered input to various drafts of the paper

